# Deep learning-derived quantitative interstitial abnormalities in early rheumatoid arthritis and healthy controls: A multicenter, prospective cross-sectional study

**DOI:** 10.64898/2026.02.20.26346723

**Authors:** Gregory C McDermott, Xiaosong Wang, Natalie A Davis, Misti L Paudel, Ying Qi, Emily Kowalski, Grace Qian, Liya S Getachew, Kevin T Mueller, Alene A Saavedra, Lauren A O’Keeffe, Madeleine Beaulé, Ritu Gill, Staci Gagne, Suzanne Byrne, Michael H. Cho, Edwin K. Silverman, Madison Negron, Kathleen MM Vanni, Caleb Bolden, Tina Mahajan, Erica Mulcaire-Jones, Neda Kortam, Paul F Dellaripa, Pierre-Antoine Juge, Tracy J Doyle, Marcy B Bolster, Kevin D Deane, Dinesh Khanna, Bryant R England, Raul San Jose Estepar, George R Washko, Jeffrey A Sparks

**Author notes:** **Corresponding Author:** Jeffrey A. Sparks, MD, MMSc Division of Rheumatology, Inflammation, and Immunity Brigham and Women’s Hospital 60 Fenwood Road, #6016U Boston, MA 02115, @jeffsparks. **Disclaimer:** The funders had no role in the decision to publish or in the preparation of this article. The content is solely the responsibility of the authors and does not necessarily represent the official views of Harvard University, its affiliated academic healthcare centres or the National Institutes of Health.

## Abstract

**Objective:** Quantitative computed tomography (QCT) can automatically quantify parenchymal abnormalities on chest CT imaging using deep learning. We leveraged QCT to detect pulmonary abnormalities in patients with early rheumatoid arthritis (RA) compared to healthy controls.

**Methods:** We analyzed high-resolution CT chest imaging from participants with early RA in the prospective, multicenter, SAIL-RA study and healthy non-smoking controls from the COPDGene study. A deep learning classifier quantified the percentage of normal lung, interstitial abnormalities, and emphysema for each participant. We compared the percentage of QCT features between early RA participants and healthy comparators and examined associations using multivariable linear regression.

**Results:** We analyzed 200 participants with early RA (median RA duration 8.3 months, mean age 55.7 years, 74.5% female) and 104 healthy controls (mean age 62.0 years, 68.3% female). The median percentage of interstitial abnormalities on QCT was 3.7% (IQR 2.1, 6.1%) for early RA and 1.6% (IQR 0.8, 2.4%) for healthy controls (p<0.0001). Early RA was associated with 9.3% less normal lung on QCT than healthy controls, adjusted for age and sex (p<0.0001). Among RA participants, QCT interstitial abnormalities were associated with older age (multivariable β=0.1 per year, 95%CI 0.07-0.2, p<0.0001) and higher DAS28-ESR (multivariable β=0.6 per unit, 95%CI 0.01-1.3, p=0.046).

**Conclusion:** Participants with early RA had less normal lung and more interstitial abnormalities on a deep learning-derived QCT measure than healthy controls. These results suggest that loss of normal lung is already present in early RA and emphasizes the urgent need for strategies to preserve lung health in RA.

## INTRODUCTION

Rheumatoid arthritis (RA) is associated with a spectrum of interstitial lung disease (ILD) severity that ranges from subclinical abnormalities to clinically apparent lung disease that contributes to significant morbidity and mortality^1–5^. Detection and monitoring of RA-ILD typically relies on visual interpretation of high-resolution computed tomography (HRCT) chest imaging, which requires significant expertise and can be insensitive to subtle abnormalities and changes over time^6^. Several quantitative CT (QCT) techniques have been developed to automatically quantify abnormal lung features using machine learning^7^. However, there has been limited application of these techniques to RA-ILD.

The early RA period represents an important opportunity to identify and quantify lung abnormalities and identify risk factors for the presence and progression of RA-ILD. Several epidemiological studies have detailed that RA-ILD can occur early in the RA disease course or prior to the development of articular symptoms^8^. Since only a subset of patients with subclinical disease will progress to clinically apparent RA-ILD, there is significant uncertainty about optimal monitoring and treatment strategies during this period. QCT techniques have been deployed in multiple research settings and have demonstrated strong associations with respiratory symptoms and all-cause mortality in population-based studies^9,10^. Additionally, QCT techniques have been used to monitor changes in interstitial abnormalities^10^ and may be an attractive approach for monitoring progression as well as the effect of immunosuppressive and antifibrotic therapies in patients with RA-ILD.

We used data from a multicenter, prospective cohort study of patients with early RA as well as a study that included healthy controls to investigate the association of QCT findings with RA and demographic, lifestyle, and RA clinical factors. All participants were assessed with HRCT chest imaging that was processed using a deep learning QCT classifier. We hypothesized that early RA would be associated with increased quantitative interstitial abnormalities compared to healthy controls and that higher RA disease activity would be associated with increased interstitial abnormalities among those with early RA.

## METHODS

### Early RA participants

We analyzed baseline data of the longitudinal prospective Study of Inflammatory Arthritis and Interstitial Lung Disease in Early RA (SAIL-RA). All participants were diagnosed with early RA, defined as within two years of study enrollment and met 2010 ACR/EULAR classification criteria for RA^11^. Study participants were recruited at five United States (US) academic healthcare centers between 2017 and 2024: Brigham and Women’s Hospital (Boston, MA), Massachusetts General Hospital (Boston, MA), University of Colorado (Aurora, CO), University of Michigan (Ann Arbor, MI), and University of Nebraska Medical Center (Omaha, NE). The SAIL-RA study was approved by the Mass General Brigham (#2020P003768) and University of Nebraska Medical Center (#0282-16-FB) Institutional Review Boards.

### Healthy controls

Healthy, never-smoker controls were recruited through the COPDGene study, a multicenter, prospective, cohort study that recruited a group of healthy never smoker comparators in addition to a larger group of current and former smokers^12,13^. COPDGene participants were recruited at 21 clinical centers in the US between 2007 and 2011. Participants completed baseline health questionnaires at enrollment that included self-reported physician diagnosis of RA and current medication use. For this analysis, we defined healthy controls as any of the healthy, never-smokers who did not self-report a history of RA or use of a disease-modifying antirheumatic drug (DMARD) medication.

This COPDGene substudy was approved by the Mass General Brigham Institutional Review Board (protocol #2020P000558). Written informed consent was obtained from all participants across both studies.

### Study measures

As previously reported^14^, all SAIL-RA participants were assessed with surveys, history, physical examination, HRCT chest imaging, pulmonary function tests (PFTs), and laboratory testing at the baseline study visit. Survey data included demographics, smoking status (never, past, or current smoker), and pack-years of cigarette smoking. HRCT scans were performed on multidetector CT scanners (at least 16 detectors) with inspiratory, supine volumetric images and image reconstruction using 1 to 1.25mm slice thickness. PFTs included forced expiratory volume in one second (FEV_1_), forced vital capacity (FVC), and diffusing capacity of the lungs for carbon monoxide (DLCO). Laboratory testing for erythrocyte sedimentation rate (ESR) was performed on peripheral blood collected during the study visit. Data on rheumatoid factor and anti-cyclic citrullinated peptide were obtained from review of clinically performed laboratory tests. Body mass index (BMI) was calculated from height and weight measured as part of the study visit physical examination. Tender and swollen joint assessments were performed by trained rheumatologists at all sites.

For each participant, current medications and RA diagnosis date were obtained from review of electronic health records. Disease Activity Score with 28 joints and ESR (DAS28-ESR) was calculated based on tender and swollen joint counts, patient global assessment, and ESR measurement performed as part of the study visit^15,16^. Disease activity was further characterized into remission or low (DAS28-ESR <3.2) or moderate or high (DAS28-ESR ≥3.2). For 11 patients who were missing ESR, we used the Clinical Disease Activity Index^16^ to determine disease activity category. Additionally, there were 13 patients missing FEV_1_ and FVC and 36 patients missing DLCO.

In COPDGene, the healthy controls provided demographic data and completed surveys at the baseline study visit. They were assessed with supine CT chest imaging performed using multi-detector CT scanners (at least 16 detector channels) with volumetric acquisitions at full inspiration as well as spirometry to obtain FVC, but DLCO was not obtained.

In SAIL-RA and COPDGene, the chest CT scans were reviewed by up to three thoracic radiologists for visual evidence of interstitial lung abnormalities according to the Fleischner criteria^17^, as previously described^14^.

### QCT analysis

For both the early RA participants and healthy controls, we analyzed their baseline chest CT scans using a previously described QCT analysis method^18^. Briefly, an ensemble of convolutional neural networks was trained to identify 2D and 3D areas of imaged lung and classify the areas of interest as normal lung, emphysema, or interstitial abnormalities using factors such as tissue density and distance from the pleural surface. Interstitial abnormalities were further subclassified into ground glass opacities, nodular changes, linear scarring, reticular changes, or honeycombing. Each feature was then summed, standardized to total lung volume, and reported a percentage of lung occupied by each feature.

### Statistical analysis

We compared baseline characteristics of the early RA participants to healthy controls using chi-squared tests for categorical variables and Wilcoxon rank-sum tests and *t*-tests for continuous variables. We compared the percentage of normal lung, interstitial abnormalities, and emphysema determined by QCT between the early RA participants and the healthy controls using Wilcoxon rank-sum tests. We also compared the percentage of interstitial abnormality subfeatures between early RA participants and healthy controls. We also compared the percentage of normal lung, interstitial lung and subfeatures, and emphysema between early RA patients with and without interstitial lung abnormalities identified by thoracic radiologists. Those indeterminate for interstitial lung abnormalities were excluded for this analysis.

Among early RA participants, we examined the correlation between the percentage of QCT features (percentage of normal lung as well as interstitial abnormalities and emphysema) with DAS28-ESR, ESR, FVC % predicted, and DLCO % predicted using Spearman’s correlation coefficients.

We assessed the association of RA with QCT features in linear regression models. We included all early RA participants and healthy comparators and determined the association of RA with QCT features in unadjusted linear regression. We then repeated our analysis after additional adjustment for age and sex. In a sensitivity analysis, we limited our analysis to the n=125 early RA never smokers and compared these to the healthy controls (all of whom were never smokers).

Among the participants with early RA, we performed unadjusted linear regression models to investigate the associations between QCT features and demographics, lifestyle factors, RA characteristics, and PFT parameters. We then performed multivariable models that assessed for associations between QCT features and significant factors identified in the unadjusted models as well as sex, given the significant associations noted between sex and RA-related lung diseases in other studies^1,19,20^. We did not include FVC or DLCO in the multivariable model since these are affected by interstitial abnormalities and thus on the causal pathway.

All analyses were performed using SAS v9.4 (Cary, NC). Two-sided p values <0.05 were considered statistically significant.

## RESULTS

### Study sample and baseline characteristics

We analyzed early RA participants (n=200) from SAIL-RA and non-RA never smokers (n=104) in COPDGene who served as the healthy controls. Baseline characteristics of these groups are described in **Table 1**. The mean age of early RA participants was 55.7 years (SD 13.9) and 62.0 years (SD 9.5) for the healthy controls. The majority of both groups – 74.5% of early RA and 68.3% of the healthy controls – were female. Both the early RA (85.0%) and healthy controls (93.3%) were predominantly White race. Of the early RA participants, 125 (62.5%) were never smokers.

**Table 1:** Baseline characteristics of participants with early RA in SAIL-RA (n=200) compared with healthy controls in COPDGene (n=104)

| Characteristic | Early RA (n=200) | Healthy controls (n=104) |
| --- | --- | --- |
| **Demographics and Lifestyle** |  |  |
| Age, years (mean, SD) | 55.7 (13.9) | 62.0 (9.5) |
| Female sex at birth | 149 (74.5%) | 71 (68.3%) |
| Race |  |  |
| Asian | 8 (4.0%) | 0 |
| Black | 10 (5.0%) | 7 (6.7%) |
| White | 170 (85.0%) | 97 (93.3%) |
| Other | 12 (6.0%) | 0 |
| Smoking status |  |  |
| Never | 125 (62.5%) | 104 (100.0%) |
| Past | 64 (32.0%) | 0 |
| Current | 11 (5.5%) | 0 |
| Smoking pack-years, median (IQR) | 0 (0, 7) | 0 |
| BMI, kg/m<sup>2</sup>, median (IQR) | 27.8 (24.3, 32.8) | 27.7 (24.6, 31.5) |
| **PFTs** |  |  |
| FEV<sub>1</sub>/FVC (median, IQR) | 0.78 (0.73, 0.82) | 0.80 (0.76, 0.84) |
| FEV<sub>1</sub>% predicted (median, IQR) | 97 (84.8, 108) | 101 (91, 113) |
| FVC% predicted (median, IQR) | 98.1 (87.5, 109) | 98 (90, 108) |
| DLCO% predicted (median, IQR) | 89 (76.6, 100) | - |
| FVC% predicted <80% | 27/192 (14.1%) | 7 (6.7%) |
| FEV<sub>1</sub>/FVC <70% | 29/193 (15.0%) | 0 |
| DLCO% predicted <75% | 38/169 (22.5%) | - |
| **RA characteristics** |  |  |
| RA duration, months (median, IQR) | 8.3 (4.1, 19.0) | - |
| DAS28-ESR (median, IQR) | 3.1 (2.0, 4.3) | - |
| TJC28 (median, IQR) | 1 (0, 4) | - |
| SJC28 (median, IQR) | 1 (0, 3) | - |
| PtGA (median, IQR) | 20 (10, 50) | - |
| ESR (median, IQR) | 16 (7, 33) | - |
| Disease activity |  |  |
| Remission/low | 107 (53.5%) | - |
| Moderate/high | 93 (46.5%) | - |
| Seropositive for anti-CCP and/or RF | 167/196 (85.2%) | - |
| Current medications |  |  |
| Glucocorticoids | 58 (29.0%) | 0 |
| Methotrexate | 141 (70.5%) | 0 |
| Other csDMARDs | 71 (35.5%) | 0 |
| TNFi | 38 (19.0%) | 0 |
| Non-TNFi b/tsDMARDs | 11 (5.5%) | 0 |

Anti-CCP = anti-cyclic citrullinated peptide antibody, b/tsDMARD = biologic or targeted synthetic disease modifying antirheumatic drug, csDMARD = conventional synthetic disease modifying antirheumatic drug, DAS28-ESR = disease activity score of 28 joints with erythrocyte sedimentation rate, DLCO = diffusion capacity of carbon monoxide, FEV<sub>1</sub> = forced expiratory volume in 1 second, FVC = forced vital capacity, HRCT = high resolution computed tomography, IQR = interquartile range, RF = rheumatoid factor, SD = standard deviation, SJC = swollen joint count, TJC = tender joint count, TNFi = tumor necrosis factor inhibitor

Among the early RA participants, the median RA duration at study baseline was 8.3 months (interquartile range - IQR 4.1 to 19.0) and 46.5% had moderate or high disease activity at baseline. Methotrexate was being used by 70.5% and 29.0% were using glucocorticoids at study baseline.

### QCT results

When comparing the QCT results between early RA participants and healthy controls in unadjusted analyses (**Table 2**), the early RA participants had less normal lung (median 90.2% vs. 96.5%, p<0.0001), more interstitial abnormalities (median 3.7% vs. 1.6%, p<0.001), and slightly more emphysematous changes (median 0.7% vs. 0.5%, p=0.01). Early RA participants also had increased median percentage of interstitial abnormality subfeatures – median 3.5% vs. 1.5% (p<0.0001) for reticular changes; 0.03 vs. 0.01% (p<0.0001) for nodular changes, and 0.1% vs. 0.03% (p<0.0001) for honeycombing when to healthy controls.

**Table 2:** Baseline QCT features of participants with early RA in SAIL-RA (n=200) compared with healthy controls in COPDGene (n=104)

| **QCT features, % (median and IQR)** | **Early RA (n=200)** | **Healthy controls (n=104)** | **p-value** |
| --- | --- | --- | --- |
| Normal | 90.2 (87.0, 92.5) | 96.5 (95.8, 97.2) | **<0.0001** |
| Interstitial | 3.7 (2.1, 6.1) | 1.6 (0.8, 2.4) | **<0.0001** |
| Reticular | 3.5 (1.9, 5.6) | 1.5 (0.8, 2.3) | **<0.0001** |
| Ground glass | 0 (0, 0.03) | 0 (0, 0) | **<0.0001** |
| Nodular | 0.03 (0.01, 0.2) | 0.01 (0, 0.03) | **<0.0001** |
| Linear scar | 0.01 (0, 0.05) | 0.00 (0, 0.01) | **<0.0001** |
| Honeycombing | 0.1 (0.07, 0.2) | 0.03 (0.02, 0.05) | **<0.0001** |
| Emphysema | 0.7 (0.3, 1.3) | 0.5 (0.3, 0.7) | **0.01** |
| Paraseptal | 0.5 (0.2, 1.0) | 0.4 (0.2, 0.5) | **0.02** |
| Centrilobular | 0.2 (0.03, 0.4) | 0.1 (0.05, 0.2) | 0.28 |

Due to rounding and small amounts of unclassified lung, values may not add to 100%.
COPDGene = genetic epidemiology of chronic obstructive pulmonary disease study, IQR = interquartile range, QCT = quantitative computed tomography, RA = rheumatoid arthritis, SAIL-RA = study of inflammatory arthritis and interstitial lung disease in early rheumatoid arthritis

When comparing early RA participants with visually-determined interstitial lung abnormalities to those without interstitial lung abnormalities, the patients with interstitial lung abnormalities had less normal lung (median 82.4% vs. 90.7%) and more quantitative interstitial abnormalities (median 11.4% vs. 3.3%) compared to those without interstitial lung abnormalities (**Supplemental Table S1**).

### Early RA vs. Healthy controls for QCT features

In the multivariable linear regression models examining the association of RA status with QCT features (**Table 3**), we observed that early RA was associated with lower percentage of normal lung (-9.3%, 95%CI -10.8 to -7.8), increased interstitial abnormalities (4.6, 95%CI 3.3 to 5.9), and increased emphysema (0.7%, 95%CI 0.3 to 1.0). We observed similar findings when we restricted the early RA participants to the subgroup of n=125 participants who were never smokers (-8.5% less normal lung in early RA vs. healthy controls; p<0.0001).

**Table 3:** Linear regression of QCT in participants with early RA in SAIL-RA (n=200) and subgroup of never smokers (n=125) compared with healthy non-smoking controls in COPDGene (n=104)

| QCT features | Adjusted* $\beta$<br>(All early RA vs<br>healthy controls)<br>(95%CI) | p-value | Adjusted* $\beta$<br>(Never smokers<br>with early RA vs<br>healthy controls)<br>(95%CI) | p-value |
| --- | --- | --- | --- | --- |
| Normal | -9.3 (-10.7, -7.8) | **<0.0001** | -8.5 (-10.0, -7.1) | **<0.0001** |
| Interstitial | 4.6 (3.2, 5.9) | **<0.0001** | 3.9 (2.7, 5.1) | **<0.0001** |
| Ground glass | 0.02 (0.003, 0.04) | **0.02** | 0.03 (0.01, 0.05) | **0.02** |
| Reticular | 4.2 (3, 5.4) | **<0.0001** | 3.6 (2.5, 4.7) | **<0.0001** |
| Nodular | 0.1 (0.06, 0.2) | **<0.0001** | 0.1 (0.05, 0.2) | **0.0007** |
| Linear scar | 0.04 (0.03, 0.06) | **<0.0001** | 0.02 (0.01, 0.04) | **0.0001** |
| Honeycombing | 0.2 (0.12, 0.2) | **<0.0001** | 0.2 (0.11, 0.2) | **<0.0001** |
| Emphysema | 0.7 (0.3, 1.0) | **0.0001** | 0.6 (0.2, 1.0) | **0.003** |
| Paraseptal | 0.4 (0.1, 0.6) | **<0.0001** | 0.3 (0.2, 0.5) | **0.0005** |
| Centrilobular | 0.3 (0.1, 0.5) | **0.003** | 0.3 (0.03, 0.5) | **0.03** |

\* = multivariable linear regression adjusted for age and sex
CI = confidence interval, COPDGene = genetic epidemiology of chronic obstructive pulmonary disease study, QCT = quantitative computed tomography, RA = rheumatoid arthritis, SAIL-RA = study of inflammatory arthritis and interstitial lung disease in early rheumatoid arthritis

### Associations with QCT interstitial abnormalities among early RA participants

We then examined correlations between QCT features and DAS28-ESR, ESR, FVC % predicted, and DLCO % predicted among the early RA participants. Results are displayed in **Figure 1** and **Supplemental Figures S1-S2**. We observed that QCT interstitial percentage was positively correlated with DAS28-ESR (rho=0.22, p=0.003) and ESR (rho=0.30, p<0.0001). Additionally, QCT interstitial percentage was inversely correlated with FVC % predicted (rho=-0.26, p=0.0003) and DLCO % predicted (rho=-0.44, p<0.0001).

**Figure 1:**
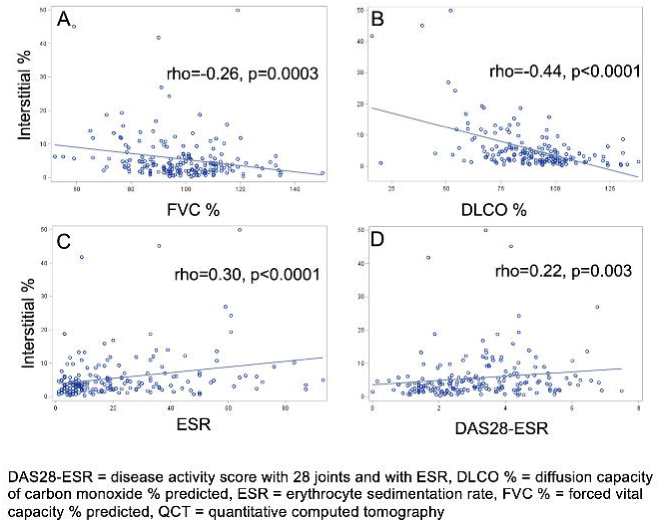
Among participants with early RA in SAIL-RA, correlations of QCT interstitial changes with (A) forced vital capacity, (B) diffusion capacity of carbon monoxide, (C) erythrocyte sedimentation rate, and (D) DAS28-ESR disease activity

Results from the linear regression models examining the associations between baseline factors with percentage of interstitial abnormalities on QCT are displayed in **Table 4**. In these models, the β represents the increase or decrease in the percentage of interstitial abnormalities associated with each factor. In the unadjusted models, older age (0.2% per year, 95%CI 0.1 to 0.2), past smoking status (+2.2% ever vs. never smoking, 95%CI 0.2 to 4.3), DAS28-ESR (+0.6 per unit, 95%CI 0.02 to 1.3), higher ESR (+0.08 per mm/hr, 95%CI 0.03 to 0.1), RF-positive status (+2.2 vs. negative 95%CI 0.001 to 4.4) were each associated with higher QCT interstitial percent. Decreasing FVC % predicted (-0.09 per unit, 95%CI -0.2 to -0.03) and DLCO (-0.2 per unit, 95%CI -0.2 to -0.1) were inversely associated with QCT interstitial percentage.

**Table 4:** Associations of baseline factors with percentage of interstitial changes on QCT among participants with early RA in SAIL-RA (n=200)

| Characteristic | Unadjusted $\beta$ | 95%CI | p-value | Multivariable* $\beta$ | 95%CI | p-value |
| --- | --- | --- | --- | --- | --- | --- |
| **Demographics** |  |  |  |  |  |  |
| Age at RA (per year) | 0.2 | (0.1, 0.2) | **<0.0001** | 0.1 | (0.07, 0.2) | **0.0001** |
| Age at RA, categorical |  |  |  |  |  |  |
| <60 years | Ref | Ref | Ref |  |  |  |
| $\geq 60$ years | 3.8 | (1.9, 5.6) | **<0.0001** | | | |
| Male (vs. female) | 1.0 | (-1.2, 3.2) | 0.36 | 0.9 | (-1.3, 3.1) | 0.43 |
| **Lifestyle** |  |  |  |  |  |  |
| Smoking status |  |  |  |  |  |  |
| Never | Ref | Ref | Ref | Ref | Ref | Ref |
| Past | 2.2 | (0.1, 4.3) | **0.04** | 0.6 | (-1.5, 2.6) | 0.58 |
| Current | -0.4 | (-4.6, 3.8) | 0.84 | -2.8 | (-7.0, 1.5) | 0.20 |
| Smoking pack-years (per unit) | -0.003 | (-0.03, 0.02) | 0.81 |  |  |  |
| BMI (per unit) | 0.06 | (-0.1, 0.2) | 0.43 |  |  |  |
| **RA characteristics** |  |  |  |  |  |  |
| DAS28-ESR (per unit) | 0.6 | (0.02, 1.3) | **0.04** | 0.6 | (0.01, 1.3) | **0.046** |
| TJC28 (per unit) | 0.02 | (-0.2, 0.2) | 0.82 |  |  |  |
| SJC28 (per unit) | 0.03 | (-0.2, 0.3) | 0.81 |  |  |  |
| PtGA (per unit) | 0.02 | (-0.02, 0.05) | 0.34 |  |  |  |
| ESR (per unit) | 0.08 | (0.03, 0.1) | **0.0006** |  |  |  |
| Moderate/high DAS28-ESR (vs. remission/low) | 3.2 | (1.3, 5.1) | **0.0009** |  |  |  |
| Seropositive (vs. seronegative) | 0.9 | (-1.9, 3.6) | 0.54 |  |  |  |
| Anti-CCP positive (vs. anti-CCP negative) | 0.7 | (-1.5, 3.0) | 0.54 |  |  |  |
| RF positive (vs. RF negative) | 2.2 | (0.001, 4.4) | **0.05** | 1.8 | (-0.4, 3.9) | 0.11 |
| Current medications |  |  |  |  |  |  |
| Glucocorticoids (vs. not) | 2.1 | (-0.03, 4.1) | 0.054 |  |  |  |
| Methotrexate (vs. not) | 0.6 | (-1.5, 2.7) | 0.58 |  |  |  |
| TNF inhibitors (vs. not) | -1.5 | (-3.9, 0.9) | 0.22 |  |  |  |
| **PFTs** |  |  |  |  |  |  |
| FEV<sub>1</sub>/FVC (per unit) | -0.1 | (-0.6, 0.3) | 0.54 |  |  |  |
| FEV<sub>1</sub>% (per unit) | -0.04 | (-0.1, 0.01) | 0.13 |  |  |  |
| FVC% (per unit) | -0.09 | (-0.1, -0.03) | **0.002** |  |  |  |
| DLCO% (per unit) | -0.2 | (-0.2, -0.1) | **<0.0001** |  |  |  |

\* = multivariable linear regression adjusted for age at RA diagnosis, sex, smoking status, RF status, and DAS28-ESR
Anti-CCP = anti-cyclic citrullinated peptide antibody, csDMARD = conventional synthetic disease modifying antirheumatic drug, DAS28-ESR = disease activity score of 28 joints with erythrocyte sedimentation rate, DLCO = diffusion capacity of carbon monoxide, FEV<sub>1</sub> = forced expiratory volume in 1 second, FVC = forced vital capacity, HRCT = high resolution computed tomography, IQR = interquartile range, MRC = modified research council, QCT = quantitative computed tomography, RF = rheumatoid factor, SD = standard deviation, SJC = swollen joint count, TJC = tender joint count, TNFi = tumor necrosis factor inhibitor

In the multivariable model that included age (per year), sex, smoking status, and DAS28-ESR (**Table 4**), older age (0.1 per year, 95%CI 0.07 to 0.2, p=0.0001) and higher DAS28-ESR (0.6 per unit, 95%CI 0.01 to 1.3, p=0.046) were significantly associated with higher interstitial percentage. We observed similar findings in the analyses examining the associations of the same baseline factors with percentage of normal lung by QCT **(Supplemental Table S2)**, with older age and higher DAS28-ESR associated with less normal lung by QCT in the multivariable model. There were no statistically significant associations between the baseline factors with QCT emphysema percentage except for older age and male sex (**Supplemental Table S3**).

## DISCUSSION

In this prospective, multicenter cohort study of participants with early RA, we found that the patients with early RA had a median of about 9% less normal lung by a QCT deep-learning measure of HRCT than healthy controls, not explained by age, sex, or smoking. This was mostly explained by excess interstitial abnormalities. We found that older age and higher RA disease activity were associated with increased interstitial abnormalities by QCT. These findings emphasize the importance of further studies investigating the use of QCT for screening and monitoring of RA-ILD and the role of inflammation in RA-ILD pathogenesis. This also suggests that normal lung architecture may be lost in pre-RA and early RA, emphasizing the need to identify strategies to preserve lung health in people with RA.

Our study adds to the prior literature investigating pulmonary abnormalities in the early RA period. In a prior study using the SAIL-RA cohort, we demonstrated that 11% of participants with early RA had subclinical ILD detected using visual inspection. We noted a strong association between subclinical ILD and moderate or high RA disease activity^14^. Another prior study of 36 Australian patients with RA, with less than 2 years of RA joint disease found that 58% had ILD-related abnormalities on a series of pulmonary tests that included HRCT, bronchoalveolar lavage, and nuclear scanning^21^. Another investigation performed HRCT in 105 Swedish patients with early untreated RA and found that 54% had some form of parenchymal lung abnormalities including nodules and interstitial abnormalities ^22^. A Japanese study of n=65 patients with less than 1 year of RA disease duration found that 13.8% had classical ILD patterns on HRCT^23^. Our findings further emphasize the significant burden of often subclinical lung findings in the early RA period.

To our knowledge, this is the first study with a specific focus on QCT findings in an early RA population while including healthy controls. One prior study examined 193 patients with RA with a median RA disease duration of 8 years and found that high attenuation areas on QCT analysis were associated with known RA-ILD risk factors including the *MUC5B* promoter variant, cigarette smoking, and higher levels of anti-cyclic citrullinated peptide antibodies but did not include healthy controls^24^. Another general population-based study performed in the MESA cohort demonstrated an association between high attenuation areas on cardiac CT and RA-associated autoantibodies, though they could not identify RA diagnosis or duration^25^. We led a prior study investigating participants with prevalent RA among the smoker group in the COPDGene study and found that RA was associated with increased quantitative interstitial abnormalities compared to non-RA smokers^26^. While those studies were largely focused on subclinical ILD, one recent study demonstrated that among patients with known RA-ILD, the percentage of interstitial QCT features was associated with mortality^27^. Notably, the ongoing FIBRONEER-SARD trial, which is investigating the use of nerandomilast in patients with RA-ILD and other forms of systemic autoimmune rheumatic disease-associated ILD is using a different QCT measure of fibrosis as a primary trial endpoint^28^.

Our study has several strengths to consider. First, we used a QCT measure that has demonstrated strong associations between quantitative abnormalities and key outcomes, including all-cause mortality in large population-based cohorts^10,29^. Second, we included healthy, non-smoking controls and demonstrated an association between RA status and QCT interstitial abnormalities, including after adjustment for age and sex and in our secondary analysis that compared never smoker early RA participants to the healthy controls. Finally, we focused on the early RA period, when early detection of subtle abnormalities may be of significant clinical and research interest.

Our study also has certain limitations to consider. First, although the healthy controls and early RA patients used the same QCT analysis protocol, the participants were recruited during different years and at different sites, which may impact our comparisons. Second, the majority of SAIL-RA participants were female and agreed to actively participate in a longitudinal research study. Thus, the generalizability of our findings to the general RA population or to more male-predominant RA populations remains unknown. Third, although quantitative interstitial abnormalities have been associated with increased mortality in population-based studies, the clinical significance of the interstitial subfeatures (e.g., “honeycombing”) is not currently known. Additionally, although we used a validated QCT method^18^, other methods such as using high attenuation areas^24,25^, CALIPER^30^, SATORI^31^, and QILD^32^ are available and may offer different results. Also, findings from the QCT-derived interstitial variable may be due to causes from other forms of lung injury that include inhalant or smoking injury, infection, and scarring so may not be due to ILD. Fourth, data on key genetic risk factors like the *MUC5B* promoter variant or proteomics markers such as Krebs von den Lungen-6 (KL-6) are not currently available for SAIL-RA participants. Finally, this analysis represents a cross-sectional analysis of the SAIL-RA cohort. Future studies to investigate the progression and impact of subclinical interstitial abnormalities are ongoing.

In conclusion, we found that participants with early RA had less normal lung and higher percentage of interstitial abnormalities detected by QCT when compared to non-RA comparators. Interstitial abnormalities in early RA correlated with pulmonary function test parameters and were associated with higher RA disease activity. These findings highlight the promise of QCT measures for detection and monitoring of RA-ILD in the early RA period and suggest that inflammation plays a central role in the genesis of ILD in early RA. Finally, these results emphasize that significant lung damage may have accrued by the time patients are diagnosed with RA and highlight the urgent need to improve disease detection methods and strategies to preserve lung health in RA.

## Data Availability

All data produced in the present study are available upon reasonable request to the authors.

