## Supplemental Table 1 for "Deep learning-derived quantitative interstitial abnormalities in early rheumatoid arthritis and healthy controls: A multicenter, prospective cross-sectional study"

**Supplemental Figure S1: Among participants with early RA in SAIL-RA, correlations of QCT normal with (A) DAS28-ESR disease activity, (B) erythrocyte sedimentation rate, (C) forced vital capacity, and (D) diffusion capacity of carbon monoxide**

**A)**

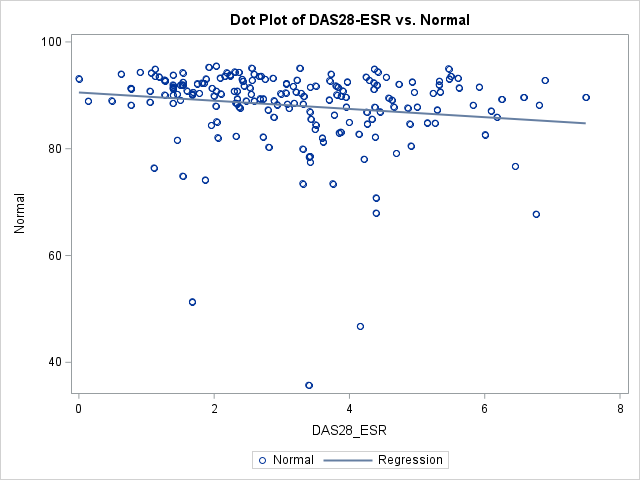
 **rho=-0.25 p=0.0007**

**B)**

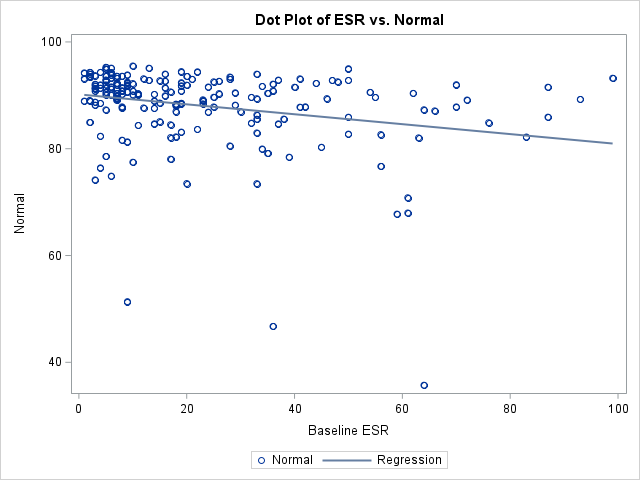
**rho=-0.32 p<0.0001**

**C)**

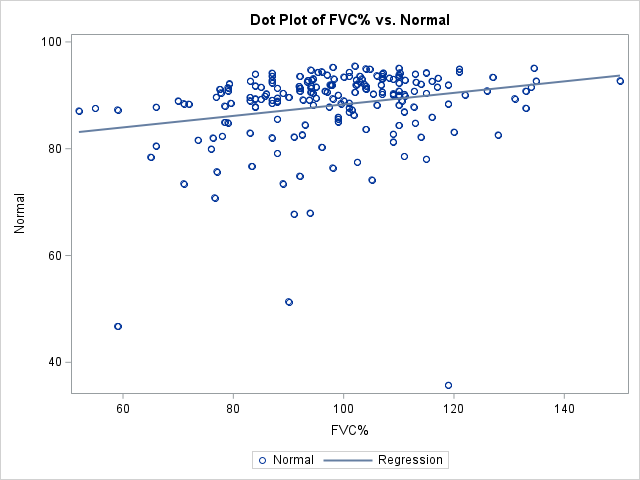
**rho=0.30 p<0.0001**

**D)**

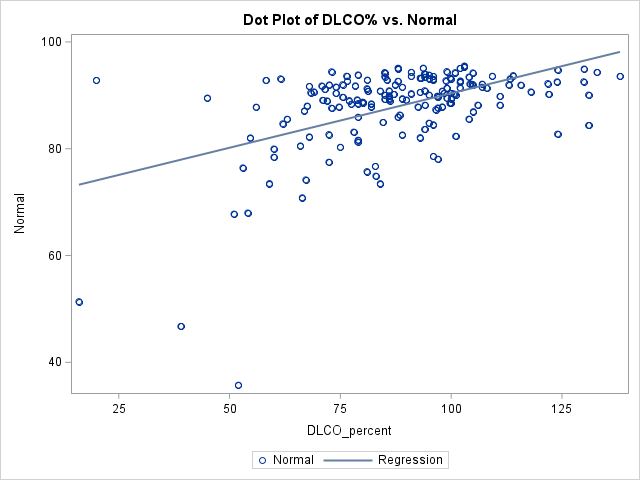
**rho=0.41 p<0.0001**

DAS28-ESR = disease activity score 28 joint with erythrocyte sedimentation rate, ESR = erythrocyte sedimentation rate, FVC = forced vital capacity % predicted, DLCO = diffusion capacity of carbon monoxide % predicted, QCT = quantitative computed tomography, RA = rheumatoid arthritis, SAIL-RA = study of inflammatory arthritis and interstitial lung disease in early RA

**Supplemental Figure S2: Among participants with early RA in SAIL-RA, correlations of QCT emphysema with (A) DAS28-ESR disease activity, (B) erythrocyte sedimentation rate, (C) forced vital capacity, and (D) diffusion capacity of carbon monoxide**

**A)**

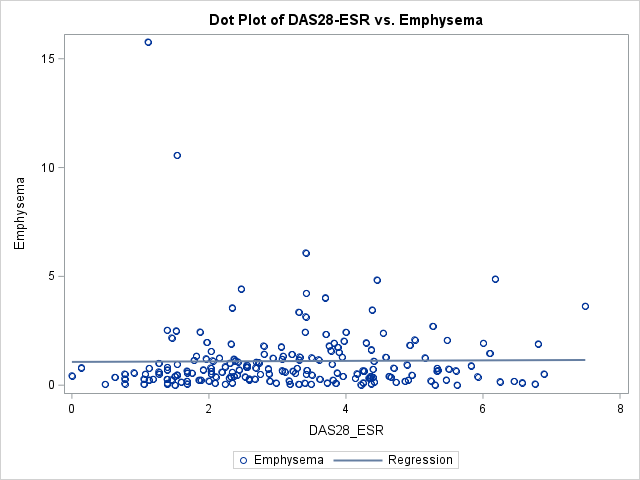
 **rho=0.11 p=0.13**

**B)**

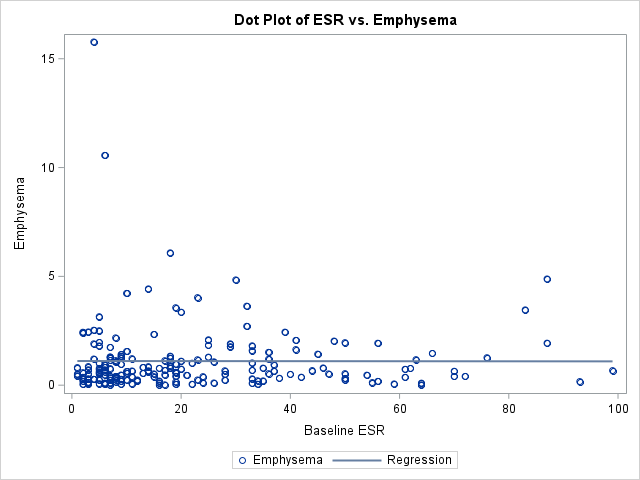
**rho=0.08 p=0.29**

**C)**

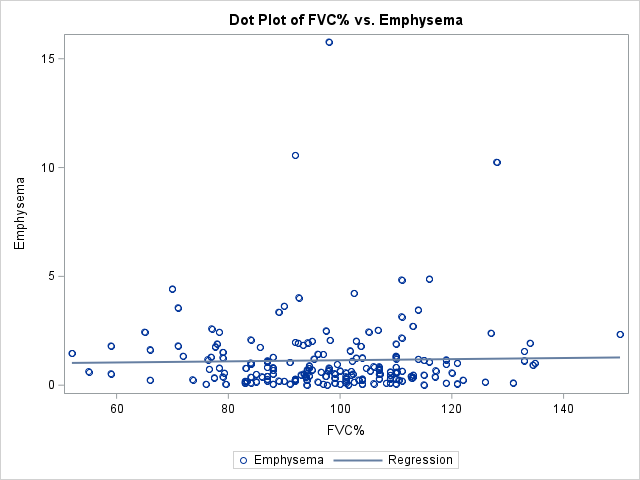
**rho=-0.02 p=0.80**

**D)**

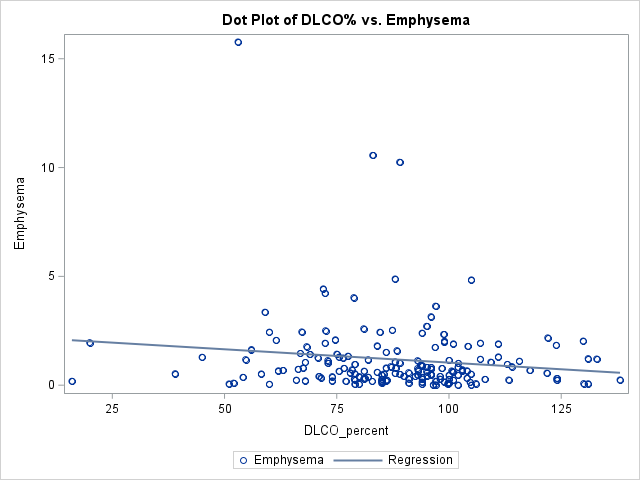
**rho=-0.11 p=0.15**

DAS28-ESR = disease activity score 28 joint with erythrocyte sedimentation rate, ESR = erythrocyte sedimentation rate, FVC = forced vital capacity % predicted, DLCO = diffusion capacity of carbon monoxide % predicted, QCT = quantitative computed tomography, RA = rheumatoid arthritis, SAIL-RA = study of inflammatory arthritis and interstitial lung disease in early RA

**Supplemental Table S1: Baseline QCT features of early participants with visually-determined interstitial lung abnormalities compared to early RA participants without visually-determined interstitial lung abnormalities (n=193)**

| **QCT features, % (median and IQR)** | **Early RA with ILA (n=26)** | **Early RA without ILA (n=167)** | **p-value** |
| --- | --- | --- | --- |
| Normal | 82.4 (73.5, 85.5) | 90.7 (88.4, 92.8) | **<.0001** |
| Interstitial | 11.4 (6.3, 18.9) | 3.3 (1.9, 5.3) | **<.0001** |
| Reticular | 11.0 (5.9, 17.8) | 3 (1.8, 4.7) | **<.0001** |
| Ground glass | 0 (0, 0.02) | 0.01 (0, 0.03) | 0.97 |
| Nodular | 0.07 (0.02, 0.4) | 0.03 (0.01, 0.1) | **0.03** |
| Linear scar | 0.1 (0.03, 0.3) | 0.01 (0, 0.03) | **<.0001** |
| Honeycombing | 0.3 (0.2, 0.7) | 0.1 (0.06, 0.2) | **<.0001** |
| Emphysema | 1.0 (0.3, 2.4) | 0.6 (0.3, 1.3) | 0.12 |
| Paraseptal | 0.7 (0.2, 1.5) | 0.5 (0.2, 0.9) | 0.16 |
| Centrilobular | 0.2 (0.1, 1.0) | 0.1 (0.02, 0.4) | **0.02** |

COPDGene = genetic epidemiology of chronic obstructive pulmonary disease study, ILA = interstitial lung abnormalities, IQR = interquartile range, QCT = quantitative computed tomography, RA = rheumatoid arthritis, SAIL-RA = study of inflammatory arthritis and interstitial lung disease in early rheumatoid arthritis

**Supplemental Table S2: Associations of baseline factors with percentage of normal on QCT among participants with early RA in SAIL-RA (n=200)**

| **Characteristic** | **Unadjusted β** | **95%CI** | **p-value** | **Multivariable* β** | **95%CI** | **p-value** |
| --- | --- | --- | --- | --- | --- | --- |
| **Demographics** |  |  |  |  |  |  |
| Age at RA (per year) | -0.21 | (-0.28, -0.14) | **<0.0001** | -0.18 | (-0.26, -0.10) | **<0.0001** |
| Age at RA, categorical |  |  |  |  |  |  |
| <60 years | Ref | Ref | Ref |  |  |  |
| ≥60 years | -4.66 | (-6.77, -2.55) | **<0.0001** |  |  |  |
| Male (vs. female) | -1.72 | (-4.22, 0.78) | 0.18 | -1.09 | (-3.57, 1.39) | 0.39 |
| **Lifestyle** |  |  |  |  |  |  |
| Smoking status |  |  |  |  |  |  |
| Never | Ref | Ref | Ref | Ref | Ref | Ref |
| Past | -2.65 | (-5.00, -0.30) | **0.027** | -0.92 | (-3.23, 1.39) | 0.43 |
| Current | 0.72 | (-4.09, 5.53) | 0.77 | 2.78 | (-1.80, 7.36) | 0.23 |
| Smoking pack-years (per unit) | 0.0005 | (-0.03, 0.03) | 0.97 |  |  |  |
| BMI (per unit) | -0.12 | (-0.27, 0.04) | 0.15 |  |  |  |
| **RA characteristics** |  |  |  |  |  |  |
| DAS28-ESR (per unit) | -0.77 | (-1.48, -0.06) | **0.0333** | -0.74 | (-1.43, -0.04) | **0.0373** |
| TJC28 (per unit) | -0.03 | (-0.26, 0.21) | 0.82 |  |  |  |
| SJC28 (per unit) | -0.04 | (-0.34, 0.25) | 0.78 |  |  |  |
| PtGA (per unit) | -0.02 | (-0.06, 0.02) | 0.29 |  |  |  |
| ESR (per unit) | -0.09 | (-0.14, -0.04) | **0.0004** |  |  |  |
| Moderate/high DAS28-ESR (vs. remission/low) | -3.72 | (-5.85, -1.59) | **0.0007** |  |  |  |
| Seropositive (vs. seronegative) | -1.11 | (-4.21, 2.00) | 0.48 |  |  |  |
| Anti-CCP positive (vs. anti-CCP negative) | -0.42 | (-3.01, 2.17) | 0.75 |  |  |  |
| RF positive (vs. RF negative) | -2.47 | (-5.01, 0.07) | 0.06 |  |  |  |
| Current medications |  |  |  |  |  |  |
| Glucocorticoids (vs. not) | -2.09 | (-4.48, 0.30) | 0.09 |  |  |  |
| Methotrexate (vs. not) | -0.45 | (-2.84, 1.95) | 0.71 |  |  |  |
| TNF inhibitors (vs. not) | 1.62 | (-1.16, 4.40) | 0.25 |  |  |  |
| **PFTs** |  |  |  |  |  |  |
| FEV_1_/FVC (per unit) | 0.13 | (-0.36, 0.63) | 0.59 |  |  |  |
| FEV_1_% (per unit) | 0.05 | (-0.004, 0.11) | 0.07 |  |  |  |
| FVC% (per unit) | 0.11 | (0.04, 0.17) | **0.00180** |  |  |  |
| DLCO% (per unit) | 0.20 | (0.15, 0.26) | **<0.0001** |  |  |  |

* = multivariable linear regression adjusted for age at RA diagnosis, sex, smoking status, and DAS28-ESR

Anti-CCP = anti-cyclic citrullinated peptide antibody, csDMARD = conventional synthetic disease modifying antirheumatic drug, DAS28-ESR = disease activity score of 28 joints with erythrocyte sedimentation rate, DLCO = diffusion capacity of carbon monoxide, FEV_1_ = forced expiratory volume in 1 second, FVC = forced vital capacity, HRCT = high resolution computed tomography, IQR = interquartile range, MRC = modified research council, RF = rheumatoid factor, SD = standard deviation, SJC = swollen joint count, TJC = tender joint count, TNFi = tumor necrosis factor inhibitor

**Supplemental Table S3: Associations of baseline factors with percentage of emphysema changes on QCT among participants with early RA in SAIL-RA (n=200)**

| **Characteristic** | **Unadjusted β** | **95%CI** | **p-value** |
| --- | --- | --- | --- |
| **Demographics** |  |  |  |
| Age at RA (per year) | 0.03 | (0.02, 0.05) | **<0.0001** |
| Age at RA, categorical |  |  |  |
| <60 years | Ref | Ref | Ref |
| ≥60 years | 0.81 | (0.33, 1.28) | **0.0010** |
| Male (vs. female) | 0.91 | (0.37, 1.45) | **0.0011** |
| **Lifestyle** |  |  |  |
| Smoking status |  |  |  |
| Never | Ref | Ref | Ref |
| Past | 0.27 | (-0.26, 0.79) | 0.32 |
| Current | 0.38 | (-0.70, 1.45) | 0.49 |
| Smoking pack-years (per unit) | 0.003 | (-0.003, 0.01) | 0.31 |
| BMI (per unit) | -0.01 | (-0.05, 0.02) | 0.53 |
| **RA characteristics** |  |  |  |
| DAS28-ESR (per unit) | 0.01 | (-0.14, 0.16) | 0.88 |
| TJC28 (per unit) | 0.003 | (-0.05, 0.06) | 0.90 |
| SJC28 (per unit) | 0.02 | (-0.04, 0.09) | 0.47 |
| PtGA (per unit) | 0.0002 | (-0.009, 0.01) | 0.96 |
| ESR (per unit) | -0.0002 | (-0.01, 0.01) | 0.98 |
| Moderate/high DAS28-ESR (vs. remission/low) | 0.07 | (-0.42, 0.55) | 0.78 |
| Seropositive (vs. seronegative) | 0.12 | (-0.57, 0.81) | 0.73 |
| Anti-CCP positive (vs. anti-CCP negative) | -0.53 | (-1.10, 0.04) | 0.07 |
| RF positive (vs. RF negative) | 0.21 | (-0.35, 0.78) | 0.46 |
| Current medications |  |  |  |
| Glucocorticoids (vs. not) | -0.05 | (-0.59, 0.48) | 0.85 |
| Methotrexate (vs. not) | 0.008 | (-0.52, 0.54) | 0.98 |
| TNF inhibitors (vs. not) | -0.31 | (-0.93, 0.30) | 0.32 |
| **PFTs** |  |  |  |
| FEV_1_/FVC (per unit) | 0.03 | (-0.08, 0.15) | 0.54 |
| FEV_1_% (per unit) | -0.01 | (-0.02, 0.002) | 0.10 |
| FVC% (per unit) | 0.003 | (-0.01, 0.02) | 0.74 |
| DLCO% (per unit) | -0.01 | (-0.03, 0.001) | 0.08 |

* = multivariable linear regression adjusted for age at RA diagnosis, sex, smoking status, RF status, and DAS28-ESR

Anti-CCP = anti-cyclic citrullinated peptide antibody, csDMARD = conventional synthetic disease modifying antirheumatic drug, DAS28-ESR = disease activity score of 28 joints with erythrocyte sedimentation rate, DLCO = diffusion capacity of carbon monoxide, FEV_1_ = forced expiratory volume in 1 second, FVC = forced vital capacity, HRCT = high resolution computed tomography, IQR = interquartile range, MRC = modified research council, QCT = quantitative computed tomography, RF = rheumatoid factor, SD = standard deviation, SJC = swollen joint count, TJC = tender joint count, TNFi = tumor necrosis factor inhibitor
